# Association between ethylene oxide exposure and Parkinson’s disease: evidence from U.S. Participants

**DOI:** 10.64898/2026.05.18.26353529

**Authors:** Huanli Zhang, Chunyan Wang, Shaojie Bi, Huanxian Liu, Wen An, Qianqian Liu

## Abstract

Ethylene oxide is a widely used industrial chemical,yet evidence linking its exposure to Parkinson’s disease remains limited.Using data from participants in the United States,we examined whether exposure to ethylene oxide is associated with Parkinson’s disease.This cross-sectional study included 8,430 adults from the National Health and Nutrition Examination Survey (NHANES) collected between 2013 and 2020.Information on demographic characteristics,socioeconomic factors,lifestyle behaviors,body mass index,sedentary time and major chronic conditions was analyzed. Levels of hemoglobin ethylene oxide adducts,a biomarker of ethylene oxide exposure, were evaluated in relation to Parkinson’s disease using statistical modeling approaches.After accounting for potential confounding factors,higher levels of ethylene oxide exposure were associated with an increased likelihood of Parkinson’s disease.The association followed a positive and linear pattern.These findings provide new population-based evidence suggesting that ethylene oxide may be linked to Parkinson’s disease and highlight the need for further studies to confirm causality and to better understand the biological mechanisms involved.

## Introduction

Parkinson’s disease (PD) is a rapidly expanding neurodegenerative disorder and is second only to Alzheimer’s disease (AD) in incidence.^1^It significantly impairs motor function in older adults and has profound adverse effects on psychological well-being and overall quality of life.^2^The primary pathological features of PD include the degeneration of dopaminergic neurons in the substantia nigra and the formation of Lewy bodies,^3^leading to characteristic motor symptoms such as resting tremor, bradykinesia,muscle rigidity,and postural instability,as well as a wide range of non-motor manifestations such as constipation,olfactory dysfunction,sleep disturbances,autonomic dysfunction,and psychiatric impairments.^4^Against the backdrop of global population aging,the prevalence of PD continues to rise. Worldwide,the prevalence is estimated at 1.51 per 1,000 individuals,increasing markedly to 9.34 per 1,000 among those aged 60 years and older.^5,6^

By 2050,the prevalence of PD is projected to reach 267 cases (230–320) per 100,000 people,representing a substantial 76% increase (range: 56%–125%) compared with 2021.As population aging accelerates globally,the number of individuals living with PD is expected to reach 25.2 million by 2050 corresponding to a 112% increase relative to 2021.^7^This dramatic rise will place a considerable economic burden on families,society,and healthcare systems,thereby posing a major global public health challenge.

Previous studies have identified age,industrialization,urbanization,genetic susceptibility,environmental contamination,infections,and exposure to pesticides and solvents as important risk factors for PD.^7,8^Ethylene oxide (EO) is a highly reactive cyclic compound widely used in industrial production,and prolonged or high-level exposure may seriously compromise human health.In the general population,EO exposure may occur during the manufacture of antifreeze,detergents,adhesives, textiles,solvents,and pesticides,as well as through medical sterilization and industrial disinfection processes.^9,10^EO can enter the body through inhalation and dermal contact; once absorbed,it reacts with hemoglobin to form the adduct N-(2-hydroxyethyl) valine,known as hemoglobin EO adducts (HbEO),which has been implicated in the development of various diseases.^11^Numerous studies have associated EO exposure with diabetes,cardiovascular disease,chronic obstructive pulmonary disease,cognitive impairment,and altered klotho levels.^12,13,14,15,16^EO is also considered neurotoxic, causing sensorimotor neuropathy,muscular weakness,hypoesthesia,and demyelinating fiber degeneration.^16,17^To date, however, no studies have examined whether EO exposure influences the risk of PD.To address this gap,we used data from the National Health and Nutrition Examination Survey (NHANES) to investigate the association between blood HbEO levels and PD among United States (U.S.) adults.This analysis aims to improve understanding of environmental risk factors for PD and to provide epidemiological evidence to support strategies for its prevention and mitigation.

## Methods

### Study population and design

The NHANES is a population-based,cross-sectional program conducted by the CDC. It employs a stratified,multistage probability sampling design to assess the health and nutritional status of the non-institutionalized civilian population in the U.S.NHANES collects comprehensive demographic and health-related information through in-home interviews and standardized physical examinations,including laboratory testing,which are conducted in mobile examination centers (MECs).The NHANES survey protocol is approved by the National Center for Health Statistics (NCHS) Research Ethics Review Board,and all participants provided written informed consent prior to participation.Because the data are de-identified and publicly available,additional institutional review board approval is not required for secondary analyses.^18^ NHANES datasets are publicly accessible at https://www.cdc.gov/nchs/nhanes/index.htm.

This cross-sectional study utilized three NHANES cycles (2013–2014, 2015–2016, and 2017–March 2020).A total of 35,706 participants were initially identified.After excluding pregnant women (*n* = 222),individuals with missing EO data (*n* = 27,048), and participants with PD’s disease information (*n* = 6),a final analytical sample of 8,430 participants was included.The participant selection process is illustrated in Figure 1.

**Fig 1.**
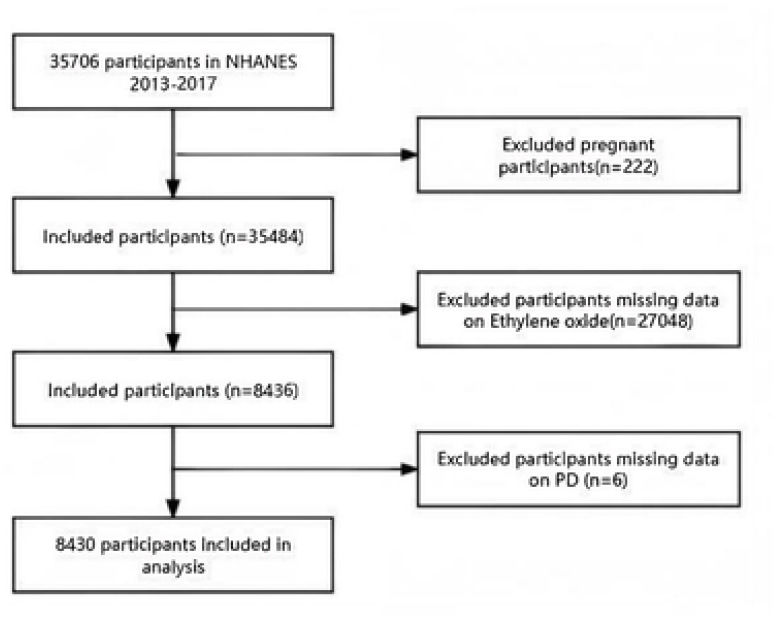
Flow chart of study participants.

### Assessment of Parkinson’s Disease

The identification of PD in the NHANES was based on participants’ responses to survey questions regarding the use of medications for PD.^19,20^Specifically,individuals who reported using medications classified as *antiparkinson agents* were defined as having PD in this study.Participants who did not report the use of such medications were categorized as not having PD.

### Measurement of ethylene oxide

Population-based data on both endogenous and exogenous exposure to EO remain limited.Hemoglobin adducts of EO,which persist in the blood for several weeks because of their relatively half-life,currently provide a sensitive biomarker for quantifying low-level exposure.^21^Measurement of HbEO followed the NHANES Laboratory Procedures Manual.Washed packed red blood cell specimens were processed and shipped to the Division of Laboratory Sciences,National Center for Environmental Health,CDC (Atlanta, GA),where they were stored at –30°C until analysis.The adducts were measured using high-performance liquid chromatography-tandem mass spectrometry (HPLC-MS/MS).For analyte concentrations below the limit of detection,imputed values were calculated as the lower limit of detection divided by the square root of two.

### Assessment of covariates

Covariates were selected based on prior references and NHANES guidelines and included sex,age,race,education level,marital status,family income,BMI,sedentary time and medical history,including hypertension,diabetes mellitus(DM),coronary heart disease(CHD),and stroke.^19,22^Race was categorized as non-Hispanic White, non-Hispanic Black,Mexican American,and other races.Marital status was classified as living with a partner or living alone.Educational attainment was grouped into < 9 years, 9–12 years, and > 12 years. BMI was calculated using standardized measurement of weight and height.Family income was categorized using the poverty-to-income ratio (PIR) as low (PIR ≤ 1.3), medium (1.3 < PIR ≤ 3.5),and high (PIR > 3.5).

Sedentary time was assessed using the NHANES questionnaire item that asked participants to report the amount of time typically spent sitting during a usual day, including time spent at school,at home,during transportation,or on leisure activities, excluding sleep.Diabetes mellitus was defined by self-reported physician diagnosis. CHD was defined by a self-reported history of congestive heart failure,coronary artery disease,angina pectoris,or myocardial infraction.Hypertension and stroke were identified based on self-reported physician diagnosis.

### Data analysis

A cross-sectional study was conducted among 8,430 U.S. participants from NHANES. The distribution of continuous variables was evaluated using histograms,Q-Q plots, and the Kolmogorov-Smirnov test.Normally distributed continuous variables are presented as means ± standard deviations,whereas skewed variables are described as medians with interquartile ranges.Group comparisons were performed using Student’s *t*-tests or Mann-Whitney *U*-tests for continuous variables and x^2^,or Fishers’ exact tests for categorical variables,as appropriate.Multiple imputation using chained equations was applied to address missing data,with three replications performed to reduce bias and preserve statistical power.Due to the skewed distribution of HbEO levels,values were natural ln-transformed and categorized into tertiles.Linear regression models were used to estimate odds ratios and 95% confidence intervals for the association between lnHbEO levels and PD.Restricted cubic spline regression with four knots (5^th^, 35^th^, 65^th^ and 95^th^ percentiles) was applied to explore potential dose-response relationships.Three models were constructed.Model 1 was adjusted for age,sex,race, education level,marital status and family income.Model 2 was further adjusted for BMI and sedentary time.Model 3 additionally included hypertension,DM,CHD,and stroke.Tests for trend were performed using multivariate regression models.Subgroup analyses were conducted according to sex,age,race,education,PIR,and BMI (< 25 kg/m^2^, 25–29.9 kg/m^2,^ and ≥ 30 kg/m^2^).

All analyses were performed using R Software (version 4.2.2) and the Free Statistics analysis platform (Version 1.9).A two-sided *P* < *0*.*05* was considered statistically significant.

## Results

### Characteristics of the population

A total of 8,430 eligible participants aged 6–80 years were included in this analysis. Baseline characteristics stratified by EO exposure are presented in Table 1,which summarizes the general characteristics of participants according to tertiles of hemoglobin EO adduct levels.Among all participants,49.7% were male,and the median age was 39 years.The overall prevalence of PD was 1.0%.In-transformed hemoglobin EO adduct levels were divided into three tertiles:Q1 (1.758–2.8888 pmol/g Hb, *n* = 2797),Q2 (2.89–3.334 pmol/g Hb, *n* = 2,819),Q3 (3.336–7.286 pmol/g Hb, *n* = 2814).Compared with participants in the lowest tertile,those in higher tertiles tended to be younger,female,non-Hispanic White,have higher educational attainment, live with a partner,have a medium PIR,have lower BMI,report no history of hypertension or DM,and have lower sedentary time (all *P* < 0.05).

**Table.1.** Characteristics of participants.

| Variables | ethylene oxide ( pmol/g Hb ) |  |  |  | *p* |
| --- | --- | --- | --- | --- | --- |
|  | Total (n = 8430) | Q1 (n = 2797) | Q2(n = 2819) | Q3 (n = 2814) |  |
| Gender n (%) |  |  |  |  | < 0.001 |
| Male | 4193 (49.7) | 1269 (45.4) | 1370 (48.6) | 1554 (55.2) |  |
| Female | 4237 (50.3) | 1528 (54.6) | 1449 (51.4) | 1260 (44.8) |  |
| Age(years), Mean ± SD | 39.0 (18.0, 60.0) | 40.0 (18.0, 61.0) | 35.0 (14.0, 59.0) | 40.0 (22.0, 59.0) | < 0.001 |
| Race/ethnicity, n (%) |  |  |  |  | < 0.001 |
| Non-Hispanic White | 2877 (34.1) | 1120 (40) | 890 (31.6) | 867 (30.8) |  |
| Non-Hispanic Black | 1915 (22.7) | 464 (16.6) | 566 (20.1) | 885 (31.4) |  |
| Mexican American | 1358 (16.1) | 494 (17.7) | 540 (19.2) | 324 (11.5) |  |
| Other Race | 2280 (27.0) | 719 (25.7) | 823 (29.2) | 738 (26.2) |  |
| Education level, n (%) |  |  |  |  | < 0.001 |
| < 9 | 827 ( 9.8) | 300 (10.7) | 286 (10.1) | 241 (8.6) |  |
| 9–12 | 3009 (35.7) | 837 (29.9) | 956 (33.9) | 1216 (43.2) |  |
| >12 | 4594 (54.5) | 1660 (59.3) | 1577 (55.9) | 1357 (48.2) |  |
| Marital status, n (%) |  |  |  |  | < 0.001 |
| Living with partner | 4922 (58.4) | 1699 (60.7) | 1726 (61.2) | 1497 (53.2) |  |
| Living alone | 3508 (41.6) | 1098 (39.3) | 1093 (38.8) | 1317 (46.8) |  |
| PIR, n (%) |  |  |  |  | < 0.001 |
| Low | 2577 (34.1) | 732 (29) | 803 (31.9) | 1042 (41.6) |  |
| Medium | 2841 (37.6) | 958 (38) | 947 (37.6) | 936 (37.4) |  |
| High | 2129 (28.2) | 834 (33) | 771 (30.6) | 524 (20.9) |  |
| BMI(kg/m<sup>2</sup>), Mean ± SD | 27.4 ± 7.8 | 28.5 ± 8.1 | 26.9 ± 7.9 | 26.7 ± 7.2 | < 0.001 |
| Hypertension, n (%) |  |  |  |  | < 0.001 |
| No | 6302 (74.8) | 2120 (75.8) | 2152 (76.3) | 2030 (72.1) |  |
| Yes | 2128 (25.2) | 677 (24.2) | 667 (23.7) | 784 (27.9) |  |
| DM, n (%) |  |  |  |  | 0.003 |
| No | 7540 (89.4) | 2543 (90.9) | 2516 (89.3) | 2481 (88.2) |  |
| Yes | 890 (10.6) | 254 (9.1) | 303 (10.7) | 333 (11.8) |  |
| CHD, n (%) |  |  |  |  | 0.374 |
| No | 7838 (93.0) | 2610 (93.3) | 2627 (93.2) | 2601 (92.4) |  |
| Yes | 592 ( 7.0) | 187 (6.7) | 192 (6.8) | 213 (7.6) |  |
| stroke, n (%) |  |  |  |  | 0.061 |
| No | 8097 (96.0) | 2698 (96.5) | 2716 (96.3) | 2683 (95.3) |  |
| Yes | 333 ( 4.0) | 99 (3.5) | 103 (3.7) | 131 (4.7) |  |
| Sedentary time, Median (IQR) | 6.0 (4.0, 8.0) | 6.0 (4.0, 9.0) | 6.0 (4.0, 9.0) | 6.0 (4.0, 8.0) | < 0.001 |
| PD, n (%) |  |  |  |  | 0.124 |
| No | 8348 (99.0) | 2770 (99) | 2799 (99.3) | 2779 (98.8) |  |
| Yes | 82 ( 1.0) | 27 (1) | 20 (0.7) | 35 (1.2) |  |

### Association between hemoglobin ethylene oxide adducts and Parkinson’s disease

The associations between HbEO levels and PD are presented in Table 2.In crude logistic regression analyses,LnHbEO levels were positively associated with PD when treated as a continuous variable (OR = 1.31, 95% CI: 1.07–1.60, *P* = 0.009).After adjustment for sociodemographic factors,including sex,age,race and ethnicity, educational level,marital status,and PIR ratio,higher LnHbEO adduct levels remained significantly associated with an increased likelihood of PD (OR = 1.28, 95% CI: 1.03–1.60, *P* = 0.025).In Model 2, which additionally adjusted for BMI and sedentary time,each one-unit increase in ln-transformed HbEO adduct levels was associated with a 29% higher odds of PD (OR = 1.29, 95% CI: 1.04–1.61, *P* = 0.023). In the fully adjusted model (Model 3),which further included hypertension, DM, CHD, and stroke,the association remained significant (OR = 1.26, 95% CI: 1.01–1.58, *P* = 0.037) for each additional unit increase in lnHbEO levels.When ln-transformed hemoglobin EO adduct levels were analyzed as tertiles,no statistically significant differences were observed between the second and third tertiles compared with the lowest tertile (all *P* > 0.05),likely due to the relatively small number of PD cases. However,the odds ratios for the highest tertile compared with the lowest tertile were consistently greater than 1 across all models (Model 1: OR = 1.44; Model 2: OR = 1.45; Model 3, OR = 1.37),suggesting a trend toward an increased risk of PD with higher levels of LnEO exposure.

**Table.2.**
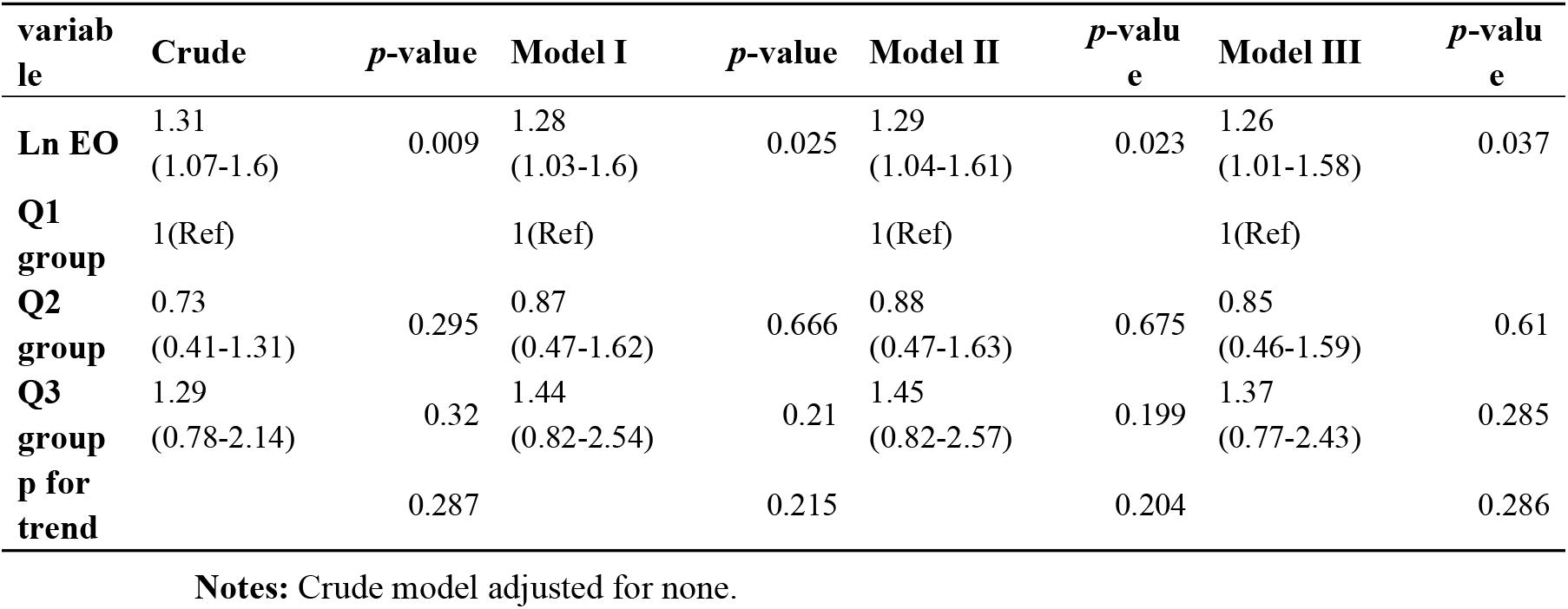
Association between ln-transformed HbEO and PD.

#### Model I

Adjust for sex,age,race and ethnicity,educational level,marital status,and PIR ratio.

#### Model II

Adjust for the variables in Model I plus BMI and sedentary time.

#### Model III

Adjust for the variables in Model II plus hypertension, DM, CHD, and stroke.

Additionally,a linear relationship between lnHbEO adduct levels and PD was observed using smooth curve fitting after adjustment for all potential confounders (*P* for nonlinearity = 0.088).The restricted cubic spline illustrating the association between lnEO and PD is presented in Supplementary Material (Fig S1).

### Subgroup Analyses

Stratified and interaction analyses were conducted to evaluate whether the association between LnHbEO and PD incidence was consistent across subgroups defined by sex, age,race,education level,PIR,marital status,and BMI (Fig. 2).Significant interaction effects were observed across age categories (*P* for interaction = 0.008) and marital status (*P* for interaction = 0.011). In contrast, no significant interactions were observed for sex, race, education, income, or BMI (all *P* for interaction > 0.05).

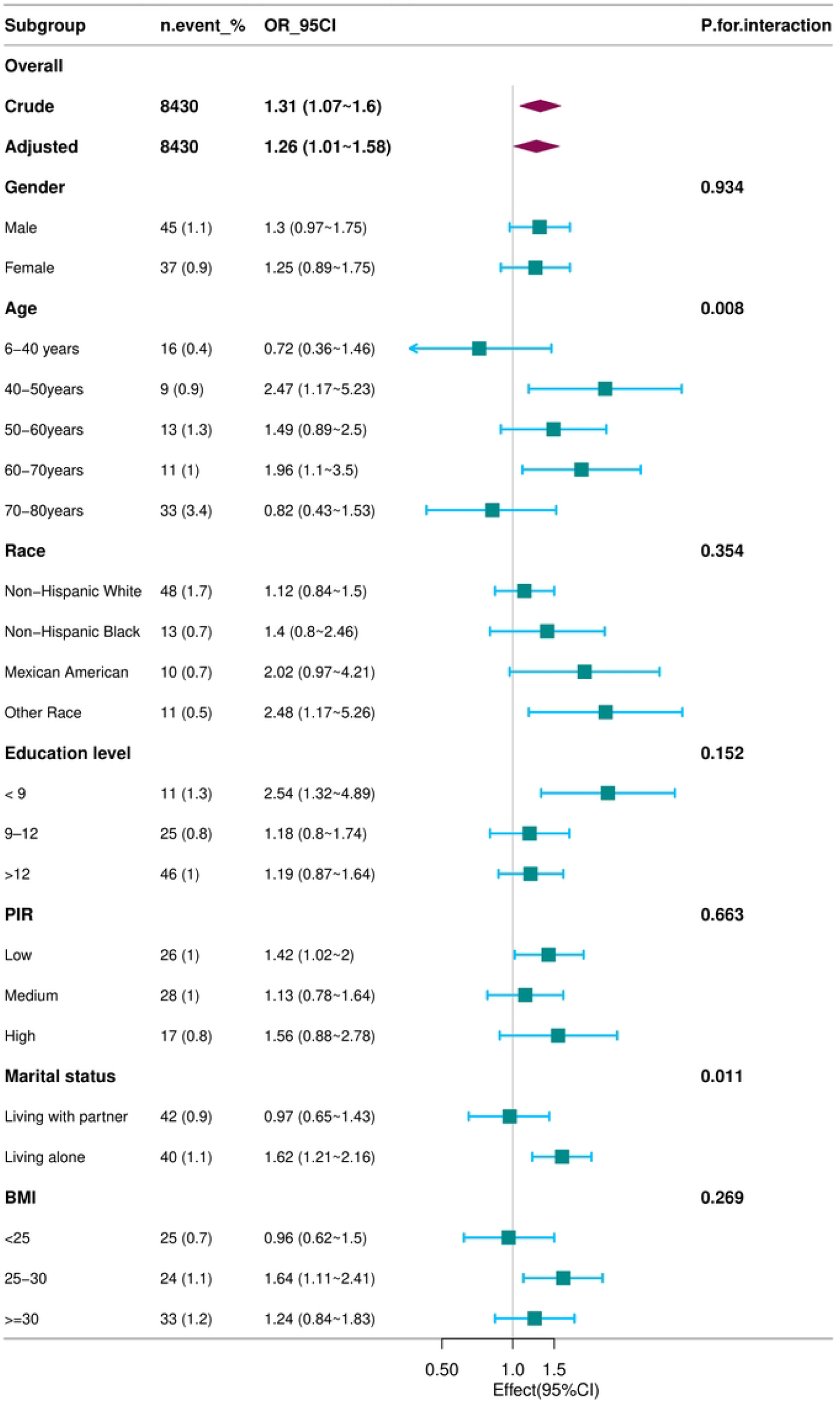

## Discussion

Using data from the NHANES database,we observed that exposure to EO was independently associated with a 26% increase in the risk of developing PD after adjusting for all covariates.In addition,restricted cubic spline analyses demonstrated a positive relationship between ln-transformed EO exposure and the incidence of PD. The global population is aging at an accelerating rate.Driven by this demographic shift,both the incidence and prevalence of PD are rising,contributing to a growing economic burden worldwide.PD is a multisystem neurodegenerative disorder characterized by the loss of dopaminergic neurons.It manifests not only as motor impairments but also as a broad spectrum of non-motor symptoms,including cognitive decline,mood disturbances,sleep disorders,autonomic dysfunction,and sensory abnormalities,^23^all of which substantially impair physical and mental well-being.PD has a multifactorial etiology involving both genetic and non-genetic components.In individuals without strong genetic risk factors,the heritability of PD is estimated to be approximately 20–30%,highlighting the substantial contribution of non-genetic factors to disease pathogenesis.^24,25^Established non-genetic risk factors include advancing age,environmental exposures,unhealthy lifestyles,metabolic disorders,and dietary factors.^26^Previous studies have reported that exposure to pesticides,air pollutants (including PM_2.5_, ozone, and NO2),and brominated flame retardants is associated with an increased risk of PD.^27,28,29,30^Barbosa and colleagues ever reported a case of parkinsonism following acute EO poisoning,^31^and our multivariable regression analysis similarly identified a positive association between EO exposure and PD.

EO is a highly reactive organic compound and a ubiquitous environmental pollutant.Exposure to EO can lead to genetic mutations and DNA damage.^32,33^A cohort study in the U.S. reported that exposure to environmental EO is associated with an increased risk of in situ breast cancer,and occupational exposure may increase mortality from lymphohematopoietic malignancies.^34^Although previous research has focused primarily on the carcinogenicity of EO,^34^recent studies in the U.S. population have demonstrated strong association with cardiovascular disease,potentially mediated by systemic inflammation and altered blood lipid profiles.^35^Elevated EO levels have also been linked to non-malignant conditions,including adolescent stroke, sleep disorders,and metabolic syndrome.^36,37,12^EO can be absorbed through the skin, directly affecting the peripheral nerves,and when inhaled,it may reach the central nervous system via vascular dissemination.^38^The biological mechanisms linking EO exposure to PD remain incompletely understood,but several pathways have been proposed.The pathophysiology of PD shares features with autoimmune disorders, involving immune and inflammatory responses.^39^Notably, oxidative stress and neuroinflammation are well-established contributors to PD pathogenesis.^40,41^Animal studies have shown that chronic EO exposure induces oxidative stress,^42^leading to alterations in Ras signaling pathways,^43^increased lipid peroxidation,disruption of the glutathione redox cycle,and reduced glutathione reductase activity.^44^These changes result in excessive production of reactive oxygen species (ROS),activation of inflammatory pathways,and upregulation of proinflammatory cytokines.^45,13^ROS damage lipids,proteins,and nucleic acids,ultimately contributing to dysfunction and loss of dopaminergic neurons,a hallmark of PD.^46^

In addition,elevated ROS in the peripheral blood of patients with PD promote platelet activation and contribute to a systemic proinflammatory state.^47^Inflammation is a well-established pathological mechanism in PD.Peripheral inflammation increases^48^ blood-brain barrier (BBB) permeability,^49^facilitating immune cell infiltration into the brain and activation of microglia in the substantia nigra,thereby promoting neurodegeneration.^50^Activation of T cells stimulates microglia and α-synuclein,a protein that can trigger both innate and adaptive immune responses. Increased expression of Toll-like receptor 4 (TLR4) in monocytes is associated with immune-mediated brain activation and altered dopaminergic neurotransmission,^51,52^ leading to sustained neuronal damage.Inflammatory responses also promote intercellular transmission of α-synuclein through an interleukin-1β/interleukin-1 receptor 1-dependentpathway,contributing to cognitive impairment and motor dysfunction.^48,53^Substantial evidence indicates that EO and its byproducts amplify oxidative stress and inflammatory responses,and chronic exposure has been shown to induce inflammatory damage across multiple organs in rodent models.^54^

Our analysis of NHANES data suggests a positive association between EO exposure and PD risk.However,several limitations should be considered.First, the cross-sectional design precludes causal inference.Second,PD was identified based on self-reported antiparkinsonian medication use,which may result in outcome misclassification;moreover,even clinical diagnosis remains presumptive because definitive confirmation requires postmortem examination.^55^Third, the relatively small number of PD cases,the absence of weighted analyses,and the potential for residual confounding warrant cautious interpretation.Despite these limitations,our findings provide population-based evidence supporting a potential link between EO exposure to PD and offer a basis for future investigations and preventative public health strategies.

## Conclusions

This study provides population-based evidence of an association between EO exposure and PD risk among U.S. adults.Prospective studies are needed to establish causality and to further elucidate the biological mechanisms underlying the association.

## Data Availability

The data in this study are publicly available from the NHANES website (http://www.cdc.gov/nchs/nhanes.htm).

## Acknowledgment

We gratefully acknowledge all participants for their valuable contributions.We also thank the Free Statistics team (Beijing, China) for providing technical support and data analysis and visualization tools.

